# The prevalence and Risk Factors Associated with Urinary Schistosomiasis among basic schoolchildren at Althawra Mobi village, Wad Madani Alkobra Locality, Gezira State, Sudan

**DOI:** 10.1101/2025.11.06.25339658

**Authors:** Taha.E Hala, Bakri Y. M. Nour

## Abstract

Schistosomiasis remains a major communicable disease of public health and socioeconomic importance in developing countries. This cross-sectional study assessed the prevalence of *Schistosoma haematobium* infection among basic schoolchildren in Al Thawra Mobi Village, Wad Madani Alkobra Locality, Gezira State, Sudan. A total of 272 participants (208 boys and 64 girls) were enrolled. Urine samples (10 ml) were collected, centrifuged at 5000 g for 5 minutes, and examined microscopically for *S. haematobium* ova; egg counts were recorded. Statistical analysis using SPSS (version 23) revealed an overall prevalence of 44% (115/261), comprising 102 mild cases and 13 severe cases. Infection was markedly higher among boys compared to girls. Risk factor analysis revealed that those children’s frequent water-contact activities around irrigation canals, such as swimming, bathing, watering animals, and washing, were significantly associated with infection. Despite tap water being the main source of drinking water, canals remained central to daily life and recreation, increasing exposure to transmission sites. The findings underscore the need for strengthened health education, improved sanitation, and integrated schistosomiasis control strategies, including preventive chemotherapy and snail control, to reduce disease burden in endemic communities.

## Introduction

Human schistosomiasis is a neglected tropical disease (NTD) of major public health and socioeconomic significance, affecting nearly 240 million people worldwide, with more than 700 million at risk of infection annually (WHO, 2023). The disease ranks among the leading causes of morbidity associated with parasitic infections, particularly in low- and middle-income countries where access to safe drinking water and adequate sanitation is limited. Approximately 91% of those requiring treatment reside in Africa, making the continent the global epicenter of schistosomiasis transmission (WHO, 2023).

Schistosomiasis is caused by blood flukes of the genus *Schistosoma*, transmitted through contact with freshwater infested with cercaria released by infected snails. Several species infect humans, with *S. haematobium* responsible for urogenital schistosomiasis (endemic in Africa and the Middle East), *S. mansoni* causing intestinal schistosomiasis (widely distributed in Africa, South America, and the Middle East), and *S. japonicum* found in East and Southeast Asia. Less common species include *S. mekongi* (Southeast Asia) and *S. intercalatum* (Central and West Africa) (CDC, 2024; Medscape, 2023).

In endemic regions, school-age children bear the greatest burden of infection due to frequent contact with infested water during daily and recreational activities. The consequences of chronic schistosomiasis include anemia, stunted growth, impaired cognition, reduced physical performance, and, in the case of *S. haematobium*, an elevated risk of bladder cancer and female genital schistosomiasis (FGS), which in turn increases susceptibility to HIV and other sexually transmitted infections (McManus et al., 2018; Christinet et al., 2016; Umbelino-Walker et al., 2023). Despite its significant health implications, FGS remains largely underdiagnosed due to limited awareness among healthcare providers in endemic areas.

Although Sudan is recognized as one of the countries most affected by schistosomiasis, up-to-date country-specific estimates of the population requiring preventive chemotherapy are limited. Earlier WHO and ESPEN reports suggested that approximately 5.7–5.8 million Sudanese required treatment in the early 2010s, representing about 15% of the national population at that time (WHO-EMRO, 2013; WHO/ESPEN, 2011). More recent figures are available at the global and regional level, with the WHO estimating that 251–265 million people required preventive chemotherapy for schistosomiasis worldwide between 2021 and 2023 (WHO, 2023). However, disaggregated figures for Sudan have not been published in accessible sources, highlighting the need for updated, transparent reporting of national preventive chemotherapy requirements to guide control and elimination strategies.

Both *S. haematobium* and *S. mansoni* are endemic, with *S. haematobium* predominating in many areas, particularly in irrigated agricultural schemes such as Gezira, New Halfa, and White Nile. Environmental and socioeconomic factors—including reliance on canal water for domestic and agricultural use, inadequate sanitation facilities, and poverty—exacerbate the persistence of transmission in these areas.

Various epidemiological studies showed that infections with *S. haematobium* and *S. mansoni* are widely distributed in different regions of Sudan. Since 1919, the disease has been first reported in northern Sudan and has been prevalent in different parts of the country, including Darfur (western Sudan), White Nile State, and Southern Kordofan State. The disease is also endemic in different villages of the Gezira agricultural scheme and New Halfa agricultural scheme.

The Gezira irrigation scheme, one of the largest in the world, has long been recognized as a hotspot for urinary schistosomiasis, where exposure is facilitated by daily human-water interactions. Previous large-scale control programs, such as the Blue Nile Health Project (1979), successfully reduced prevalence through a combination of mass drug administration (MDA), snail control, improved sanitation, and health education. However, the disease has resurged in many communities due to population growth, migration, limited sustainability of interventions, and continued dependence on canal water for daily activities. The Gezira Agricultural irrigated sachem overwhelmed by the high prevalence rates of infection (M. Amin and H. Abubaker, 2017).

It is important to note that these prevalence figures were reported prior to the outbreak of the recent war in Sudan, and the current epidemiological situation may be further exacerbated by conflict-related disruptions in health services and water management.

Gezira state is an irrigated scheme, lies between latitudes (13-32 and 15-30) North and longitudes (22-32 and 20-34) East. Khartoum State to the North, Sinnar State to the South, Gadarif State to the East and White Nile State to the West (fig 1) border it.

**Fig (1).**
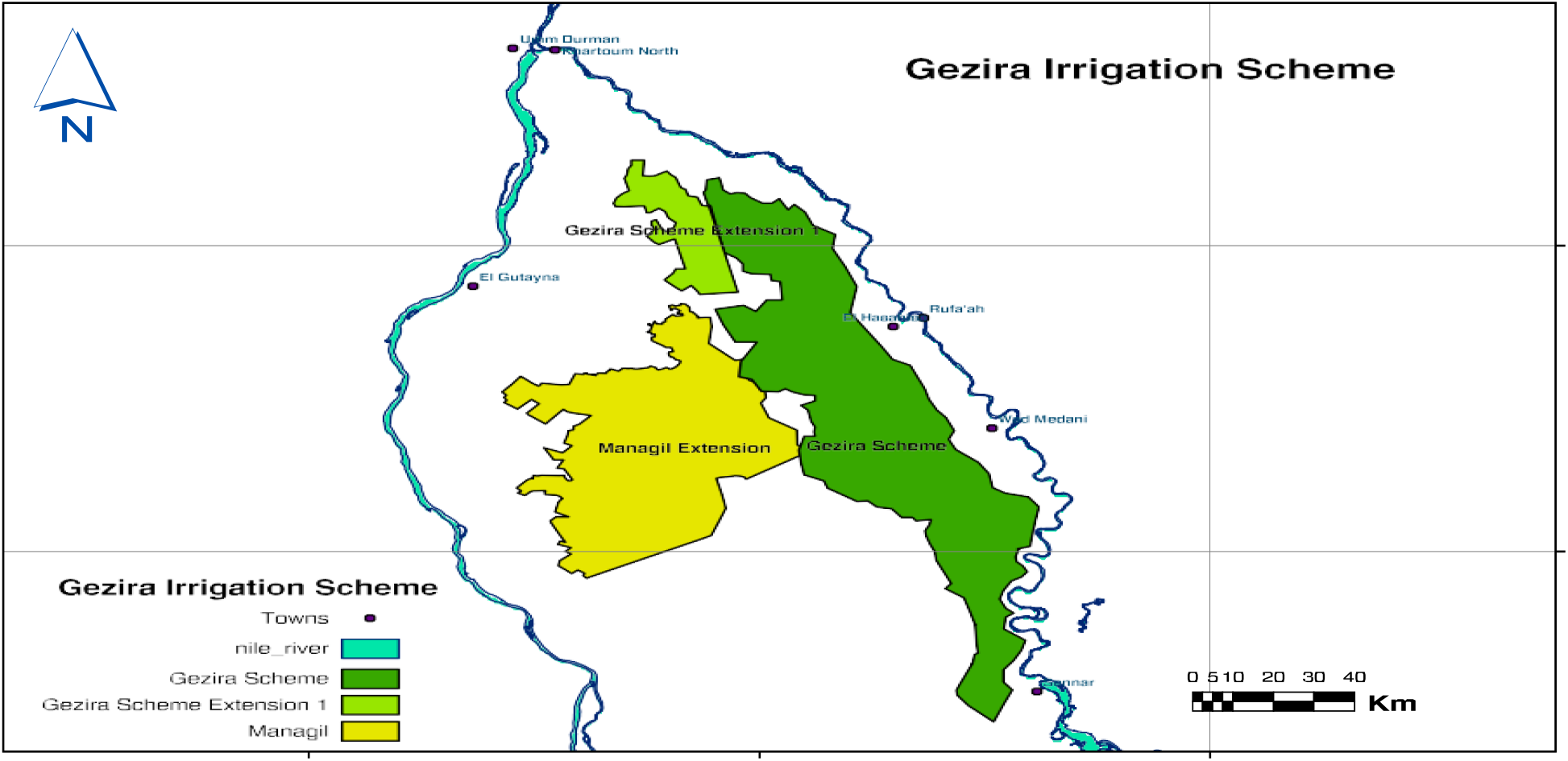
Map of the Gezira Irrigation Scheme, Sudan.

### The Blue Nile Project

In 1979, the Blue Nile Health Project (BNHP) was established to control malaria, schistosomiasis and diarrheal diseases in the Gezira-Managil and Rahad Schemes. The objective was to reduce the overall prevalence of S. mansoni and S. haematobium to less than 10%. A comprehensive approach, including treating infected people, snail control, improving sanitation and water supply, providing health education and encouraging community participation through educating schoolchildren and tenants to protect themselves and their families, was implemented. In many respects, the BNHP was a great success story in the history of the control of water-associated diseases. The overall prevalence of schistosomiasis was reduced from 53% to 6%. It should be noted that the outcome of the studies conducted by the BNHP on various aspects of schistosomiasis such as the focality of transmission, aquatic vegetation in the Gezira irrigation canals and community participation were added values for the improvement of control of schistosomiasis, in not only Sudan but worldwide (M. Amin and H. Abubaker, 2017).

The present study was designed to determine the prevalence and intensity of urinary schistosomiasis among basic schoolchildren in Al Thawra Mobi Village, Wad Madani Alkobra Locality, Gezira State, Sudan, and to identify key sociodemographic and environmental risk factors contributing to ongoing transmission. By generating updated local epidemiological data, this study aims to support national control efforts and inform strategies for more effective disease prevention and management.

The study was conducted simultaneously with a Mass Drug Administration program round targeting schoolchildren and dealing with Schistosomiasis and soil-transmitted helminth infections, which was the second round mediated by the program, were the first one was conducted on 2017.

Our study aims at conducting a descriptive Epidemiological study in order to determine the prevalence of urinary Schistosomiasis among schoolchildren and the risk factors associated with Urinary Schistosomiasis infection among basic school children in Al thawra Mobi Village located in wad madani alkobra locality, Wad Madani, Sudan.

### Study area

The study was conducted in Althawra mobi village, wad madani alkobra locality, Gezira state, it is located South-West Wad Medani city and bordered by two major canals (Fig 2). The water source in this village mainly is tab water. The canal is another source of water for nomads (living in temporary huts), field workers but it is subject to contamination by human activities, and animals at the same time, furthermore, human waste find their way to the canal since proper latrines are not accessible to some villagers specially the nomads and field workers.

**Fig (2).**
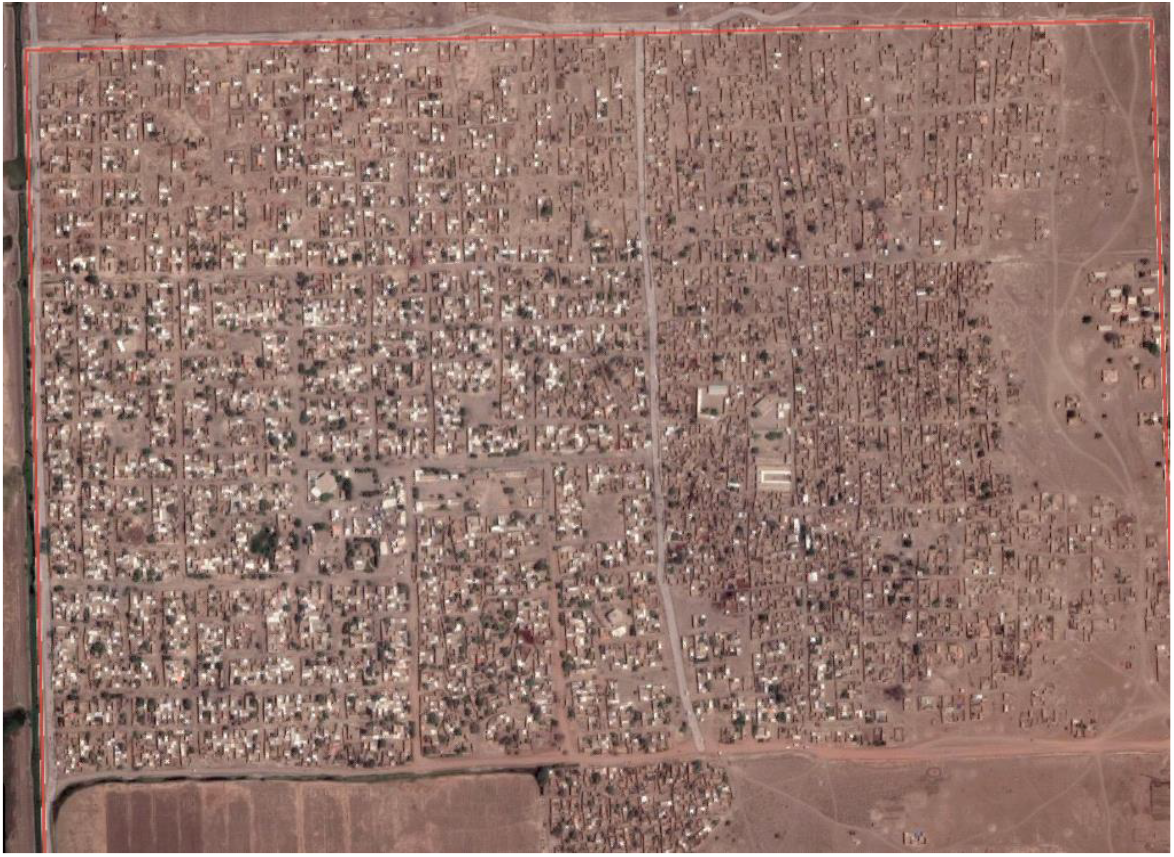
google map picture for Althawra Mobi Village

**Figure (3).**
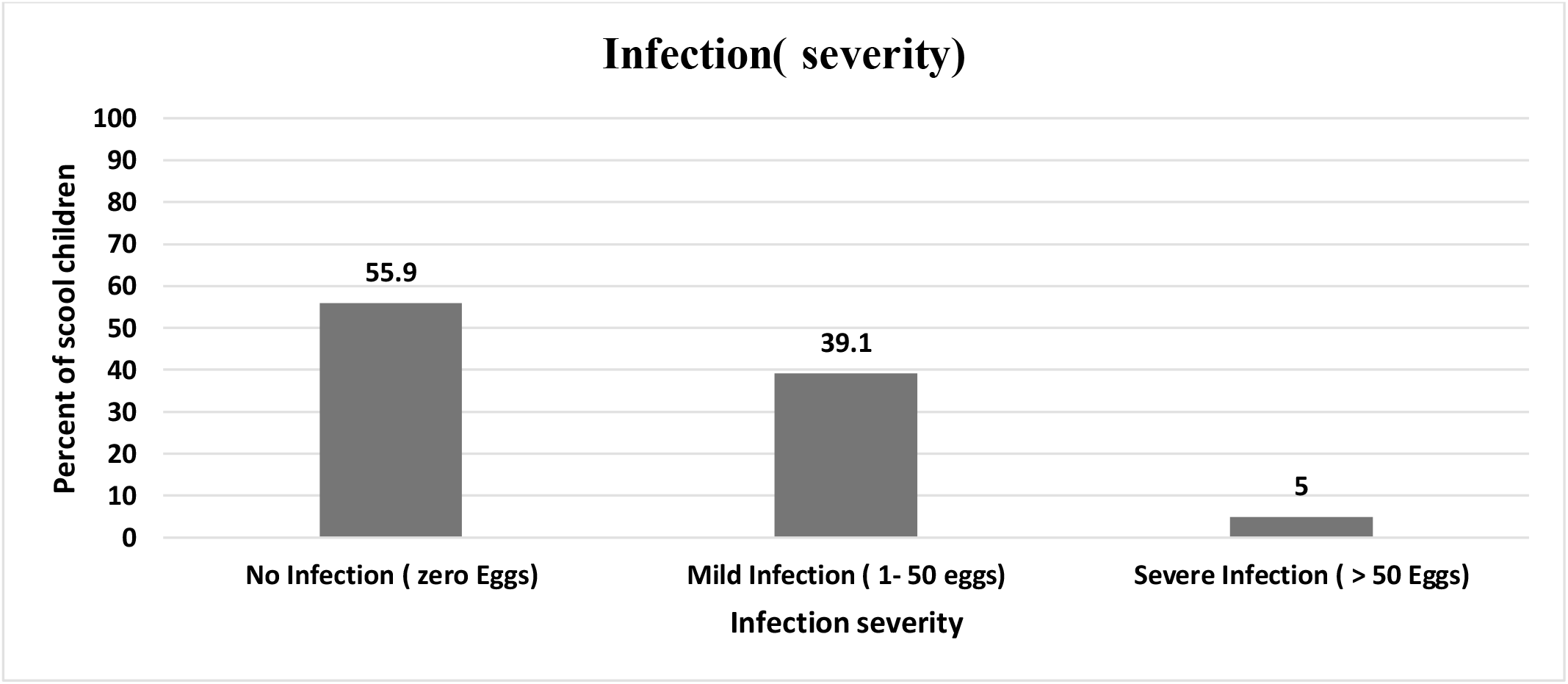
intensity of Schistosomiasis Infection among school children (n=261)

### Study population

The total population in Althawra Mobi village is estimated to be 9553 according to the National Statistics Center at Wad Madani. The study was conducted on primary school children; there are four basic schools, two for each gender. Two primary schools in this heavily infected agricultural irrigated scheme were selected, Mubarak Gasm Allah for boys and the other counterpart for girls. The two schools are closely neighbored. Samples were collected using simple random sampling of classes from the first class to the eighth class, to reach the needed study sample size, which was calculated to be 263 pupils, 208 boys, and 55 girls. Participants had been informed that they could withdraw from the study without any consequences. Written and signed or thumb-printed informed consents were obtained from schoolchildren.

### Inclusion criteria

Eligibility for inclusion into this study included:

1. Aged ≥ 5-16 years old at recruitment
2. Production of a urine sample
3. Parental/guardian consent to participate in the study.

### Exclusion Criteria

4. Students who had received PZQ in the last 6 months were excluded from the study
5. Participants who had existing medical conditions were excluded from the study.
6. Children vomiting within 4 hours after drug administration would be excluded from the study.

These criteria were based on the World Health Organization (WHO) Manual of Preventive Chemotherapy.

School health administration, the state ministry of health, and the ministry of education were contacted for permission to visit the schools, collect urine specimens, and treat children. The results of the study were shared with both parties. Principles of the participating schools explained the purpose of the survey and the expected benefits for the children and the community. A simple random sampling method was used to select participants.

### Study Design

A cross sectional descriptive study was conducted to determine the prevalence of Urinary Schistosomiasis in selected basic schools in the study area.

## Material and methods

### Data collection

Data on socio-demographic and risk factors were obtained using structured questionnaires.

### Sample collection

Urine samples were collected from all the children enrolled in the study in clean-labelled wide mouthed urine containers with lids, between 11a.m. to 1:00 p.m. terminal urine sample was collected after short exercise. Urine containers were distributed on the day of the survey. The collection team in the presence of school principals organized children, data were collected from each child, and each child was given a separate container for urine collection. The labelled capped containers containing the urine samples were transported with a cool box to the Blue Nile National Institute for Communicable Diseases for examination.

### Microscopic examination

10 ml of fresh urine were collected in clean containers with corresponding labels. Samples were processed by centrifugation at 5000g for 5 min and microscopically examined for the presence of ova of S.H, and eggs for each samples were counted. Number of counts were recorded. The intensity of infection would be categorized according to the WHO classification as negative for no detectable eggs; light for 1–49 eggs/10 ml urine; or heavy for > 50 eggs/10 ml urine.

### Treatment

Infected children’s were treated by Praziquental were it is the treatment of choice for Urinary Schistosomiasis; Children’s were weighed (according to WHO protocol) and treated with 600mg/kg. No adverse events were encountered within 24 hours after treatment

## Results

### Prevalence and severity of Schistosomiasis

Prevalence of Schistosomiasis among schoolchildren was 44% (115/261); 102 cases were mild

### Schistosomiasis rate of infection among males and females

Infection among males was 96.5% while females indicated 3.5% of infection (fig 4).

**Figure (4).**
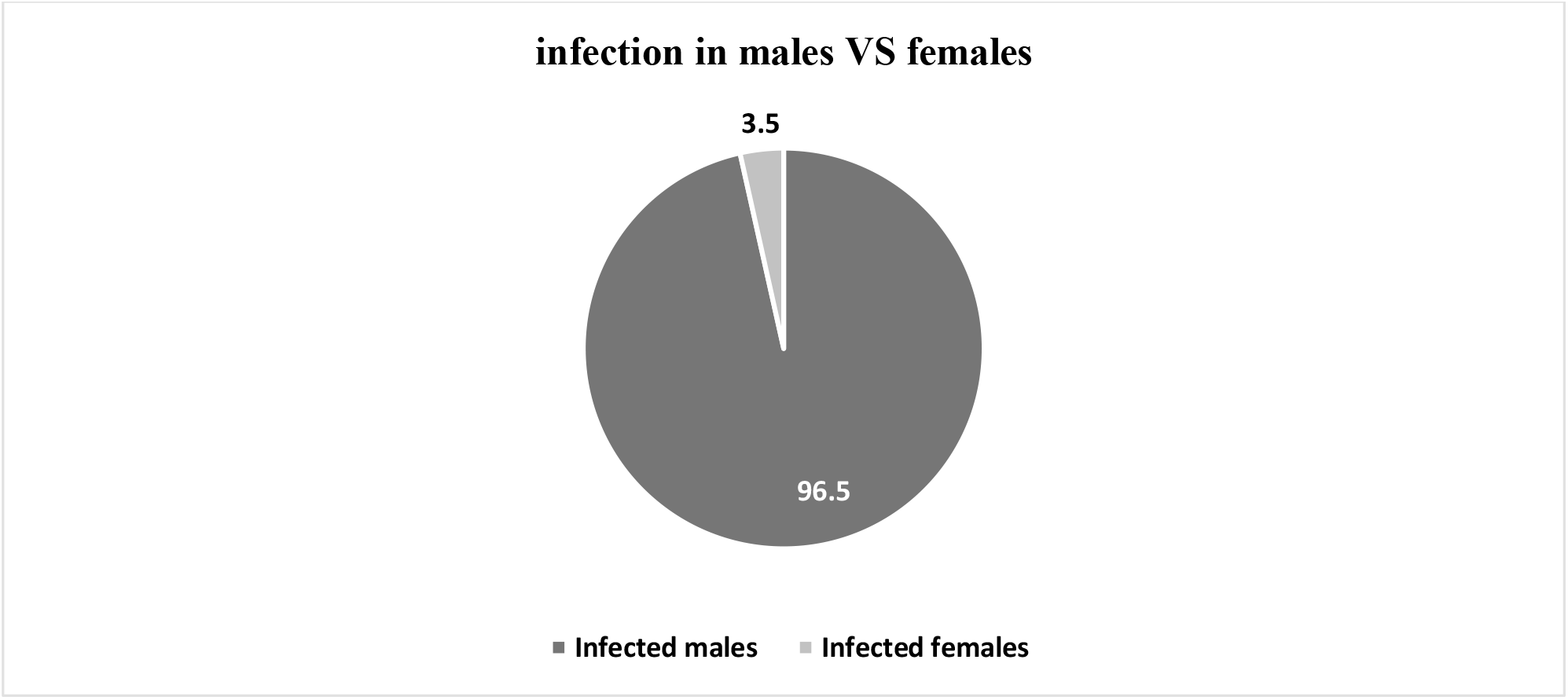
Schistosomiasis rate of infection among males and females (n=115)

### Risk factors associated with Schistosomiasis infection

Multiple factors are associated with Schistosomiasis infection. Source of water contributes directly to infection, the majority of people use tap water as the main water source, alternatively, hand water pumps are used in a minor way. Less people use river water for daily activities (fig 5). Children’s life is based essentially around the canal were multiple activities are performed there, from bathing, swimming, animal drinking, washing clothes and cars. (Fig 6) indicate these activities. Defecation practice is affected by the presence of proper latrines both at home and school. Latrines are present at school but put it is insufficient, lack water.

**Figure (5).**
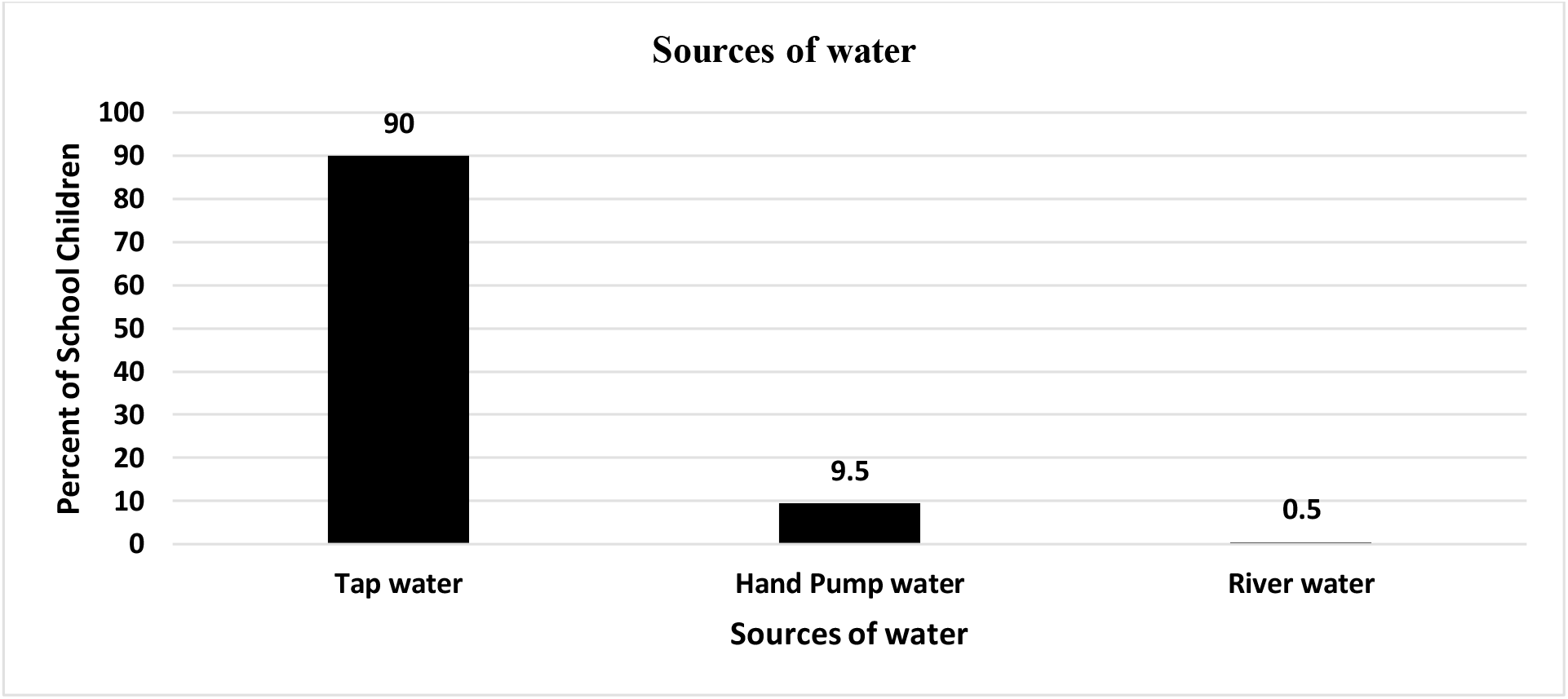
Sources of water reported by the School Children (n=221)

### 1-Sources of water

The main source of water is tap water; hand pump water and using river water also exist.

## Discussion

In this survey conducted on basic schoolchildren, a 44% prevalence is relatively high. The geographic localization of the village, surrounded by two major canals, may explain the high prevalence, as multiple activities are being conducted around the canals.

Increased prevalence of infection among males may be explained by behavioral and cultural aspects, where boys are allowed to move in a free manner to canals (precisely boys up to ten years old) where they perform different activities such as swimming or fishing in infested water, washing cars, and other activities, making them especially vulnerable to infection. In contrast, girls (especially young girls) are not allowed or given this right; they can move only with their parents or relatives. This could be explained from a safety perspective.

Our study revealed that swimming and bathing are the most prominent activities, followed by animal drinking, and finally washing clothes and cars. It is clear that the canal acts as a social and exercise club, children play and have fun there, especially in the summer time. All these activities constitute direct contact with fresh running water rich in snails, which are the main vector for the parasite (*Bulinus* sp. and *Biomphalaria* sp.).

Despite the fact that tap water is the main water source in the village, core activities are centralized around the canal. It is the major gathering point in the village.

Knowledge of the disease and its transmission upon analyzing the questionnaire showed a severe lack of information and gaps in knowledge. Although some of the participants believed that transmission could be due to canal water, others believed that transmission could be due to the sun, soil, and even hot paper (data not shown).

### Recommendations

We emphasize the importance of control of schistosomiasis, which is based on large-scale treatment of the at-risk population group. Although S. haematobium was shown to be more sensitive to praziquantel than S. mansoni, the drug showed moderate efficacy in areas that had received multiple MDA. It is recommended to conduct follow-up in endemic areas after mass drug administration to prevent possible resistance to praziquantel, where some reports have revealed reduced sensitivity to the drug.

We recommend the importance of hygiene and health education to fill the gap in knowledge among pupils and inhabitants of the village. We also recommend having more access to clean water and improving sanitation.

In parallel, we recommend the importance of conducting vector control programs via the eradication of snails abundant in the canal using multiple methods, including (but not limited to) biological eradication and chemical eradication.

## Data Availability

All data produced in the present study are available upon reasonable request to the authors

## Notes

### Competing Interest Statement

The authors have declared no competing interest.

### Funding Statement

This study did not receive any funding

### Author Declarations

Ethics Committee of Blue Nile National Institute for Communicable Diseases, University of Gezira gave ethical approval for this work (Approval No. 1/ت/خ/44).

## References

1 WHO Fact Sheet Schistosomiasis. Feb 1, 2023. [(accessed on 01 October 2024)]. Available online: https://www.who.int/news-room/fact-sheets/detail/schistosomiasis.

2 Centers for Disease Control and Prevention (CDC). “DPDx - Schistosomiasis Infection.” https://www.cdc.gov/dpdx/schistosomiasis/index.html

3 “Schistosomiasis (Bilharzia): Background, Pathophysiology, Etiology.” Medscape, https://emedicine.medscape.com/article/228392-overview

4 McManus, D. P., Dunne, D. W., Sacko, M., Utzinger, J., Vennervald, B. J., & Zhou, X. N. (2018). Schistosomiasis. Nature Reviews Disease Primers, 4(1), 13. DOI: 10.1038/s41572-018-0013-8.

5 Christinet, V., Lazdins-Helds, J. K., Stothard, J. R., & Reinhard-Rupp, J. (2016). Female genital Schistosomiasis (FGS): From case reports to a call for concerted action against this neglected gynecological disease. International Journal for Parasitology, 46(7), 395–404.

6 Umbelino-Walker, I., Wong, F., Cassolato, M., Pantelias, A., Jacobson, J., & Kalume, C. (2023). Integration of female genital schistosomiasis into HIV/sexual and reproductive health and rights and neglected tropical diseases programmes and services: A scoping review. Sexual and Reproductive Health Matters, 31(1), 2262882. 10.1080/26410397.2023.2262882

7 WHO-EMRO. (2013). Working together to eliminate schistosomiasis. World Health Organization – Regional Office for the Eastern Mediterranean. Retrieved from https://www.emro.who.int/sdn/sudan-events/launch2013-schistosomiasis.htm

8 WHO/ESPEN. (2011). The Sudan and South Sudan. Schistosomiasis Control Initiative /WHO. Retrieved from https://schisto.stanford.edu/pdf/Sudan%20and%20South%20Sudan.pdf

9 World Health Organization. (2023). Schistosomiasis (Bilharzia) – Preventive Chemotherapy Data Portal. Geneva: World Health Organization. Retrieved from https://apps.who.int/neglected_diseases/ntddata/sch/sch.htm

